# Clinical and molecular diagnostic study of 320 Chinese children with epilepsy by Genome Sequencing

**DOI:** 10.1101/2020.07.16.20153254

**Authors:** Dongfang Zou, Lin Wang, Jianxiang Liao, Hongdou Xiao, Jing Duan, Tongda Zhang, Zhenzhen Yin, Jing Zhou, Haisheng Yan, Yushan Huang, Jianbiao Li, Nianji Zhan, Ying Yang, Jingyu Ye, Fang Chen, Shida Zhu, Feiqiu Wen, Jian Guo

**Affiliations:** Department of Neurology, Shenzhen Children’s Hospital, Shenzhen, China; BGI-Shenzhen, Shenzhen 518083, China; Department of Hematology and Oncology, Shenzhen Children’s Hospital, Shenzhen, China

**Keywords:** epilepsy, Genome Sequencing, Copy Number Variant, seizure, Single Nucleotide Variations

## Abstract

**Purpose:** To evaluate the diagnostic value of Genome Sequencing(GS)in children with epilepsy.

**Methods:** We performed GS on 320 Chinese children with epilepsy and interpreted Single Nucleotide Variants (SNVs) and Copy Number Variant (CNVs) of all samples. The complete pedigree and clinical data of the probands were established and followed up. The clinical phenotypes, treatments, prognoses and genotypes of the patients were analyzed.

**Results:** Pathogenic/likely pathogenic variants were found in 122 of 320 children (38.13%), of whom, 92 (28.8%) had SNVs, 27 (8.4%) had CNVs, and three had both SNVs and CNVs. Among these variants, there were 3 CNVs of <100K in length. The most frequently mutated gene was *SCN1A*(10.9%,10/92),which is related to Dravet Syndrome, followed by *PRRT2*(8.7%,8/92), which is relevant to benign familial infantile epilepsy, *TSC2*(7.6%,7/92), which is associated with Tuberous Sclerosis. The most common recurrent CNVs were 17p13.3 deletion (18.5%, 5/27), 16p11.2 deletion syndrome (14.8%, 4/27), 15q11.2 deletion (11.1%, 3/27), which are related to epilepsy, developmental retardation and congenital abnormalities.

The diagnostic yield was higher as the age of seizure onset was smaller. The highest detection rate was 75% in whom developed seizures within one month after birth. 13.4% (43/320) cases were identified to be treatable based on GS. 1% (3/320) of epilepsy patients received direct therapeutic measures and demonstrated favorable prognosis.

**Conclusion:** GS can complete the genetic diagnosis, individualized treatment, and family reproductive guidance for patients. GS can replace Exome Sequencing and Chromosomal Microarray Analysis and is expected to be the first choice of genetic testing method for patients with epilepsy.

## Introduction

Epilepsy is one of the most common nervous system diseases. According to the World Health Organization (WHO), there are about 50 million people suffering from epilepsy in the world, which is one of the most prevalent neurological disease in childhood. Epilepsy is regarded as a group of diseases characterized by recurrent seizures.

The causes of epilepsy include the following categories: structural, genetic, infectious, metabolic, immune and unknown causes. Although epilepsy can be caused by many underlying causes, the etiology of the disease remains unknown in about 50 percent of total cases [1].

There are a number of pathogenic variations in epilepsy, for example, Copy Number Variants (CNVs), Single- Nucleotide Variants (SNVs), Small insertions/deletions (Indels), and repeat expansions. Many epilepsy genes are associated with more than one typical phenotype. Variants in the same gene can lead to different epileptic syndromes, and the same epilepsy syndrome may also be related with different genes [2]. Since the identification of the first epilepsy gene in 1995 [3], the number of genes associated with epilepsy [4] and the types of genetic tests have significantly increased. Since most of the patients with epilepsy presented with good prognosis with appropriate treatment, it is important to explore the molecular genetic mechanisms of epilepsy, so as to provide evidence for definitive diagnosis, accurate treatment and genetic counseling.

Patients with epilepsy can be tested by Next Generation Sequencing (NGS) techniques, for example capture- based targeted sequencing, Exome Sequencing (ES) and Genome Sequencing (GS) [5]. Compared with capture- based targeted sequencing and ES, GS has some superiorities. First, GS can sequence the entire genome of an individual including SNVs, small fragment insertions and deletions, and CNVs, and is superior to Chromosomal Microarray Analysis (CMA) in terms of the identifiable CNV size[6]. However, capture-based targeted sequencing missed identification of unknown genes and ES missed deep intronic SNVs, such as 20N of *SCN1A* [7], non-coding repeat expansions in familial adult myoclonic epilepsy[8], SNVs in noncoding RNA, small and large structural variants, et al. Second, library preparation for GS is simpler and faster than capture-based targeted sequencing and ES. GS shows the potential of a single test with the ability to capture almost all genomic variations in an unbiased manner, due to even coverage throughout the genome because of very limited DNA amplification and no capture step for enrichment[9].

However, previous studies have paid less attention on evaluating the diagnostic value of GS in epilepsy especially for children with epilepsy. To fill this gap, our study performed assessed the diagnostic yield and clinical utility of GS in 320 Chinese children patients with epilepsy

## Method

### Participant recruitment and eligibility criteria

We recruited unrelated patients with epilepsy under 18 years from the Department of Neurology of Shenzhen Children’s Hospital (Shenzhen, China) from October 2016 to December 2017. The diagnosis of epilepsy was made by pediatric neurologists based on the seizure semiology, electroencephalography and brain imaging results, etc.

The inclusion criteria were:

1. Diagnosis of epilepsy based on International League Against Epilepsy criteria(ILAE2017)[10].
2. Age <18 years, who were suspected to be related with genetic causes.

The exclusion criteria were:

1. Acquired brain injury such as: head trauma, brain tumors, central nervous system infections, cerebrovascular pathological changes, etc.
2. Patients after transplant surgery, stem cell therapy, transfusion of allogeneic blood products within six months, or with hemolytic disease.

We collected 2-4 ml of whole blood from 320 children of epilepsy and their parents. Genomic DNA was extracted from peripheral blood with the QIAamp DNA Blood Mini Kit (Qiagen). DNA from the patients and their parents were collected. The patient samples were sequenced by GS, and the parental samples were used for family verification with sanger verification. Pedigrees were collected and clinical data for probands were obtained through interviews and followed up.

Our research has been approved by the Institutional Review Board On Bioethics and Biosafety of BGI and the ethics committee of the Shenzhen Children’s Hospital. Written informed consent was obtained from all parents or legal guardians of the patients.

### Genome Sequencing (GS)

GS were performed on the patients with epilepsy in BGI-Shenzhen by BGISEQ-500 platform performing paired-end (PE) 100 sequencing as previously described[11].

All samples were analyzed for SNVs and CNVs. Deep sequencing data were aligned to the reference GRCh Build 37 (hg19) and variants were called using the Edico Dragen analysis pipeline[12]. Edico Genome’s Dragen Bio-IT Platform was based on the company’s Dragen Bio-IT Processor, a bioinformatics application-specific integrated reference-based mapping, aligning, sorting, deduplication, and variant calling.

CNV calls were generated using SpeedSeq SV pipeline[13](v.0.0.3a), which uses Lumpy[14] (v.0.2.9) to process samples and SVtyper[13] (v.0.0.2) to genotype variants. The pipeline output deletions, duplications, inversions, and breakpoints that cannot be assigned to one of the three classes. Low coverage data was analyzed based on pipeline[15] which calculated karyotype and ploidy based on the ratio of CNVs of cases compared to reference genomes.

Finally, variants were annotated using bcfanno(v1.4) in Frequency data(ExAC[16], GnomAD[17], G1000[18]), gene- disease data(ClinVar[19], CGD[20], OMIM[21], HGMD[22]) and other database. All of the filtered variants were then classified to seven categories based on their clinical significance (pathogenic, likely pathogenic, carry pathogenic, carry likely pathogenic, uncertain significance, likely benign, and benign) according to the American College of Medical Genetics and Genomics(ACMG) Guidelines Revisions[23, 24], and we referred to pathogenic variants and likely pathogenic variants as Diagnostic ‘positive variants’. Sanger sequencing was performed to validate the putative pathogenic variants and for parental segregation analyses where possible.

In our study, the ACMG guidelines were used to interpret the SNV results[25]. The interpretation of CNV was performed using our own laboratory interpretation rules(Figure S1), and was reviewed using ACMG’s latest CNV interpretation criteria[24]. Meanwhile, Pediatric neurologists with extensive clinical experience combined these results with the patient’s phenotype and disease progression to interpret the final genetic diagnosis. The plot of diagnostic SNVs and diagnostic CNVs on chromosomes were drawn using the R package ‘RIdeogram’[26].

### Analytical Performance of GS and ES

Because of limited DNA amplification and no capture step for enrichment, GS has easier workflow for laboratories and more uniform depth of sequencing coverage than ES. We selected thirteen GS sample data and seven ES data in our study for comparison as shown in Table S1. For GS average coverage was 42.18×, with a minimum of 99.04% of each genome covered to at least 10-fold. For ES average coverage was 166.59×, with 96.72% of target bases covered to at least 10-fold.

In the coding sequence (CDS) targeted by ES capture, the mean number of SNVs detected were 14,162, whereas it was 14,856 for GS. We then compared ES and GS for the detection of insertions/deletions (indels); the mean number of indels detected was 393 with ES and 470 with GS. For both SNVs, transitions, transversions and indels, detection rates were higher for GS than for ES. Given the greater number of SNVs, transitions, transversions and indels, more uniform depth of sequencing coverage and more reliable CNV detection, we use GS to detect 320 epilepsy patients.

### Statistical Analysis

A chi-square test of independence was performed using IBM SPSS Statistics version 19 (SPSS 19.0 (IBM, Chicago, USA).) to evaluate the diagnostic yield across different patient groups, which was the proportion of children with pathogenic/ likely pathogenic variants. For magnetic resonance imaging (MRI) findings group comparisons, patients with undetermined magnetic resonance imaging findings were excluded. P < .05 was considered statistically significant.

## Result

### Demographics and clinical data

A total of 320 Chinese children were recruited. The demographic, clinical and neuroimaging characteristics are shown in Table 1. The ratio of male to female was 1.44:1. Age at testing ranged from 1 day to 17 years, with median age was 4.3 years.

**Table 1.**
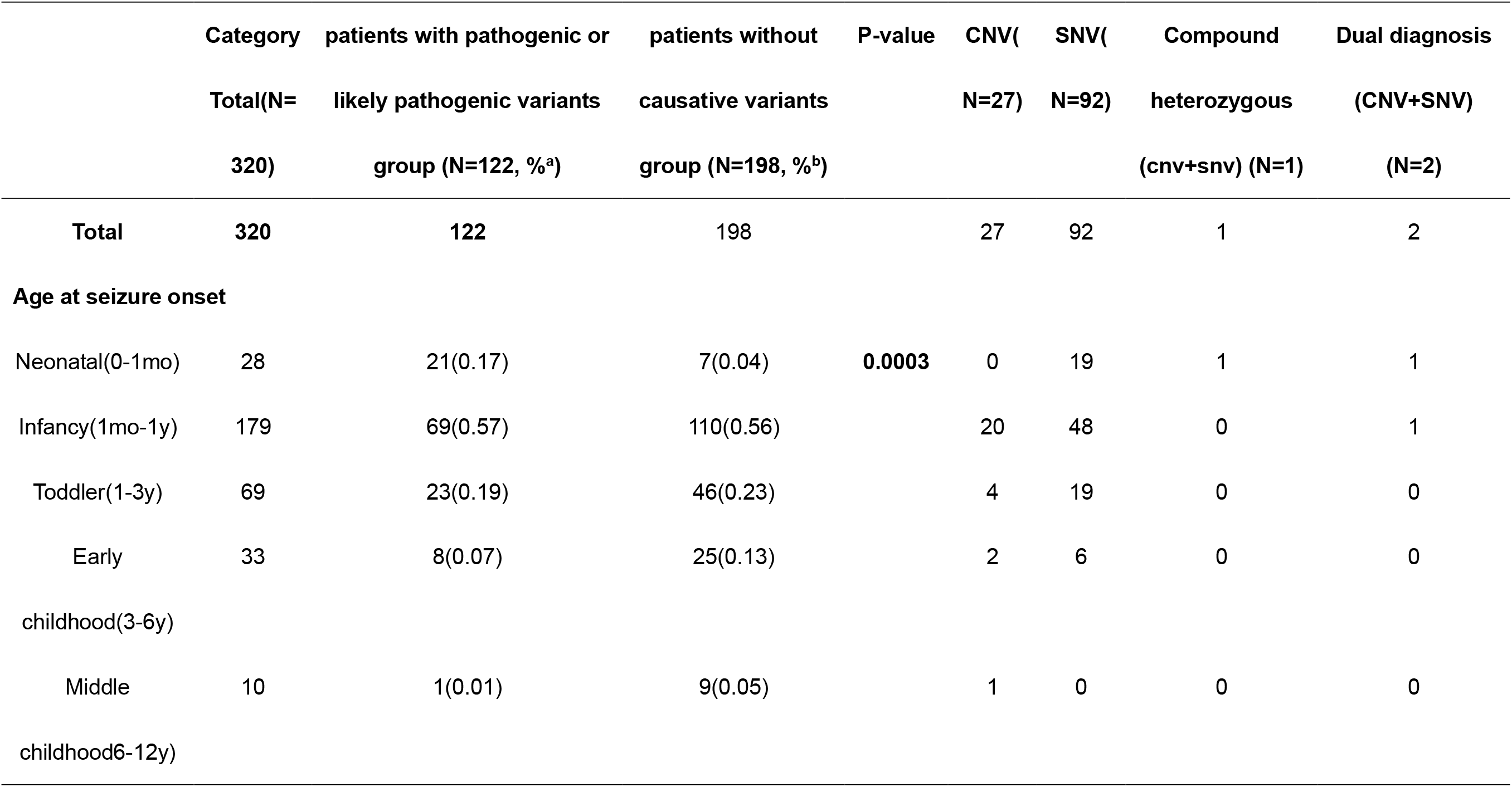
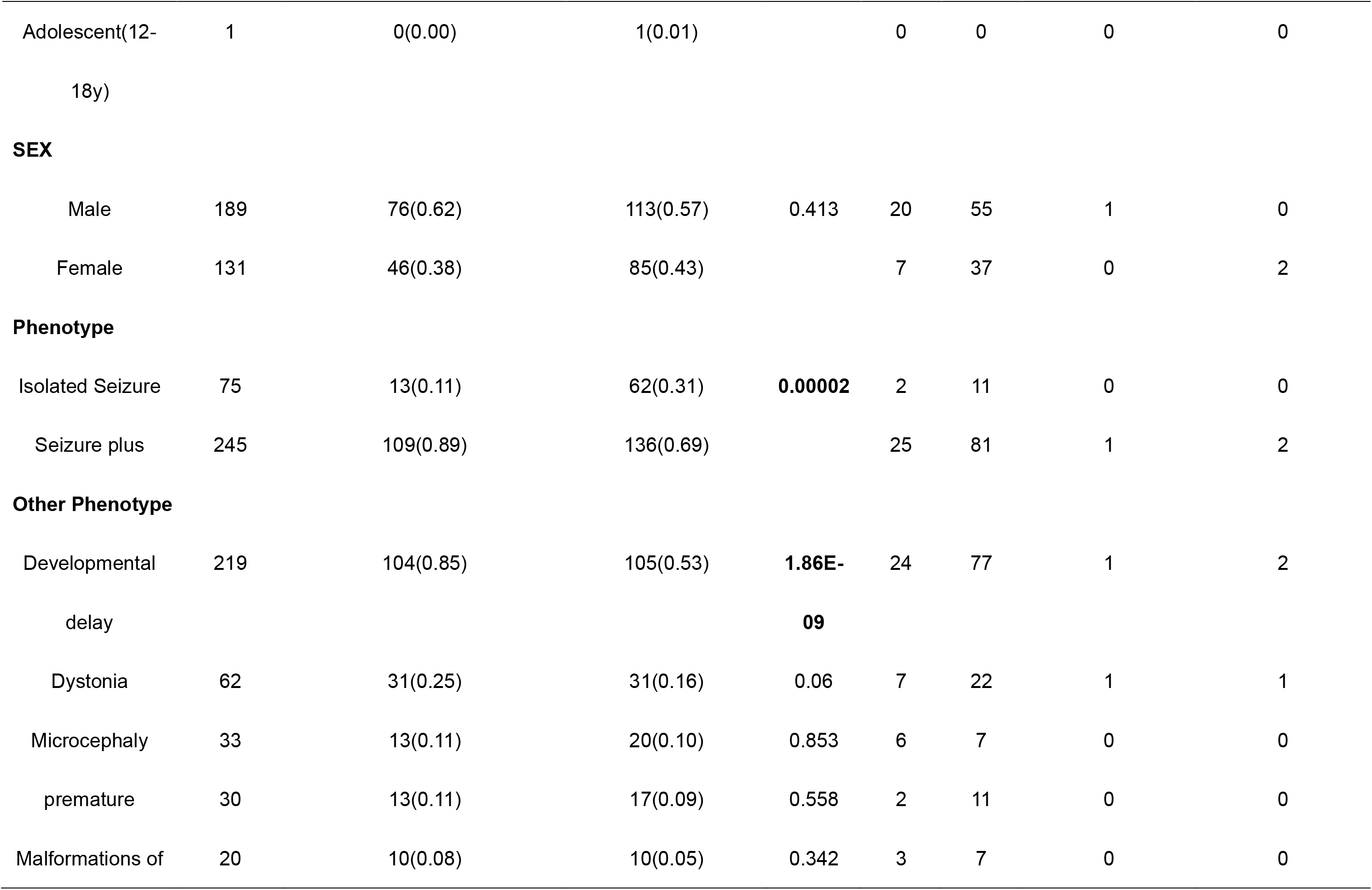
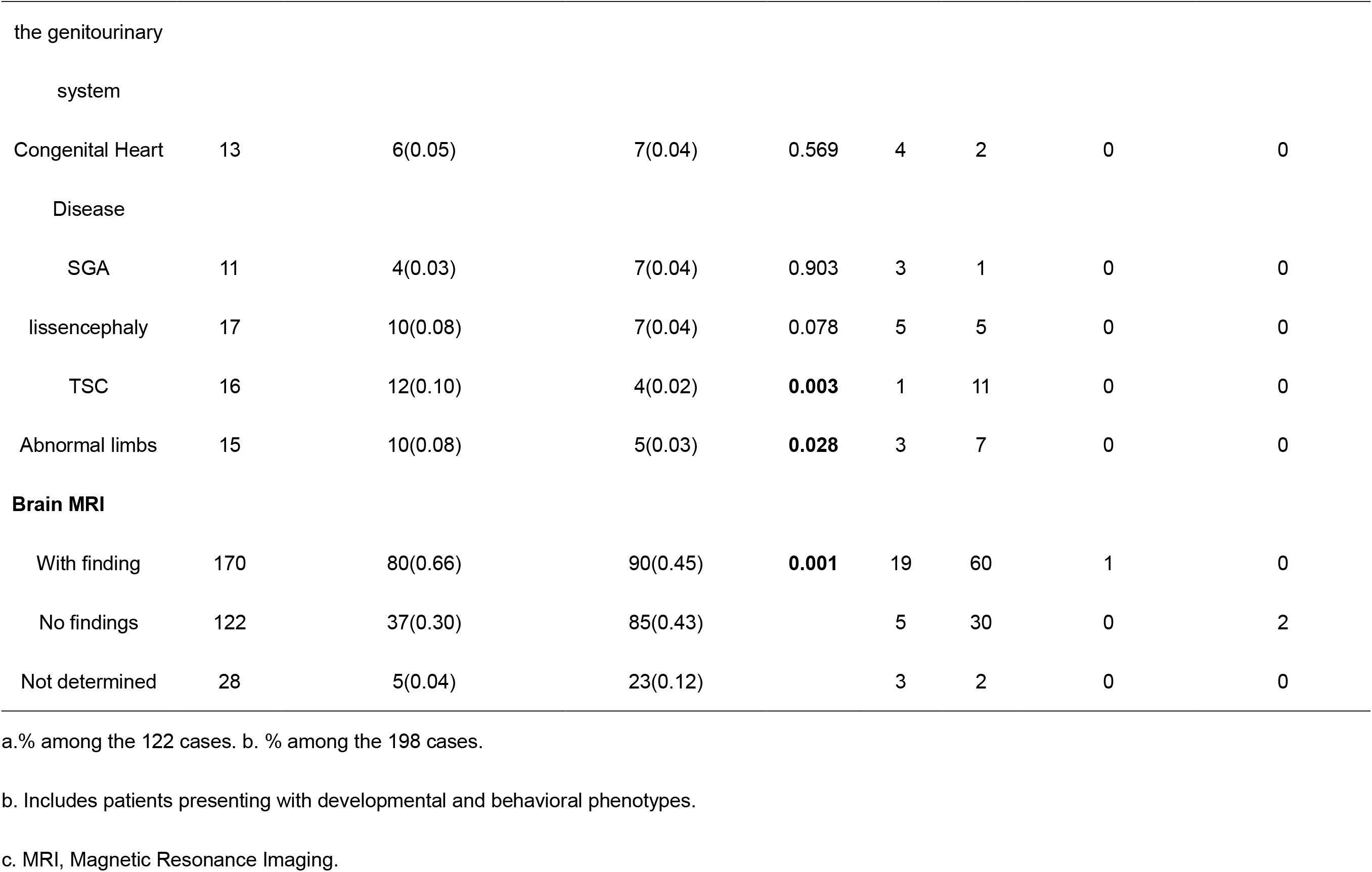
**Comparison of general clinical information in patients with pathogenic or likely pathogenic variants and the patients without causative variants**

|  | Category | patients with pathogenic or | patients without | P-value | CNV( | SNV( | Compound | Dual diagnosis |
| --- | --- | --- | --- | --- | --- | --- | --- | --- |
|  | Total(N= | likely pathogenic variants | causative variants |  | N=27) | N=92) | heterozygous | (CNV+SNV) |
|  | 320) | group (N=122, %<sup>a</sup>) | group (N=198, %<sup>b</sup>) |  |  |  | (cnv+snv) (N=1) | (N=2) |
| **Total** | **320** | **122** | 198 |  | 27 | 92 | 1 | 2 |
| **Age at seizure onset** |  |  |  |  |  |  |  |  |
| Neonatal(0-1mo) | 28 | 21(0.17) | 7(0.04) | **0.0003** | 0 | 19 | 1 | 1 |
| Infancy(1mo-1y) | 179 | 69(0.57) | 110(0.56) |  | 20 | 48 | 0 | 1 |
| Toddler(1-3y) | 69 | 23(0.19) | 46(0.23) |  | 4 | 19 | 0 | 0 |
| Early | 33 | 8(0.07) | 25(0.13) |  | 2 | 6 | 0 | 0 |
| childhood(3-6y) |  |  |  |  |  |  |  |  |
| Middle | 10 | 1(0.01) | 9(0.05) |  | 1 | 0 | 0 | 0 |
| childhood6-12y) |  |  |  |  |  |  |  |  |
| Adolescent(12-18y) | 1 | 0(0.00) | 1(0.01) |  | 0 | 0 | 0 | 0 |
| **SEX** |  |  |  |  |  |  |  |  |
| Male | 189 | 76(0.62) | 113(0.57) | 0.413 | 20 | 55 | 1 | 0 |
| Female | 131 | 46(0.38) | 85(0.43) |  | 7 | 37 | 0 | 2 |
| **Phenotype** |  |  |  |  |  |  |  |  |
| Isolated Seizure | 75 | 13(0.11) | 62(0.31) | **0.00002** | 2 | 11 | 0 | 0 |
| Seizure plus | 245 | 109(0.89) | 136(0.69) |  | 25 | 81 | 1 | 2 |
| **Other Phenotype** |  |  |  |  |  |  |  |  |
| Developmental delay | 219 | 104(0.85) | 105(0.53) | **1.86E-09** | 24 | 77 | 1 | 2 |
| Dystonia | 62 | 31(0.25) | 31(0.16) | 0.06 | 7 | 22 | 1 | 1 |
| Microcephaly | 33 | 13(0.11) | 20(0.10) | 0.853 | 6 | 7 | 0 | 0 |
| premature | 30 | 13(0.11) | 17(0.09) | 0.558 | 2 | 11 | 0 | 0 |
| Malformations of | 20 | 10(0.08) | 10(0.05) | 0.342 | 3 | 7 | 0 | 0 |
| the genitourinary |  |  |  |  |  |  |  |  |
| system |  |  |  |  |  |  |  |  |
| Congenital Heart Disease | 13 | 6(0.05) | 7(0.04) | 0.569 | 4 | 2 | 0 | 0 |
| SGA | 11 | 4(0.03) | 7(0.04) | 0.903 | 3 | 1 | 0 | 0 |
| lissencephaly | 17 | 10(0.08) | 7(0.04) | 0.078 | 5 | 5 | 0 | 0 |
| TSC | 16 | 12(0.10) | 4(0.02) | **0.003** | 1 | 11 | 0 | 0 |
| Abnormal limbs | 15 | 10(0.08) | 5(0.03) | **0.028** | 3 | 7 | 0 | 0 |
| **Brain MRI** |  |  |  |  |  |  |  |  |
| With finding | 170 | 80(0.66) | 90(0.45) | **0.001** | 19 | 60 | 1 | 0 |
| No findings | 122 | 37(0.30) | 85(0.43) |  | 5 | 30 | 0 | 2 |
| Not determined | 28 | 5(0.04) | 23(0.12) |  | 3 | 2 | 0 | 0 |

a.% among the 122 cases. b. % among the 198 cases.
b. Includes patients presenting with developmental and behavioral phenotypes.
c. MRI, Magnetic Resonance Imaging.

Among these 320 patients as shown in Figure S2a, focal seizures(68.4%,219/320) were the most common seizure type, followed by infantile spasms (31.3%,100/320), generalized tonic seizures (9.7%,31/320), myoclonic seizures (8.1%,26/320), absence seizures (3.1%,10/320), generalized tonic-clonic seizures (1.3%,4/320), and generalized clonic seizure (0.3%,1/320). There were two or more types of clinical seizure in 30% of patients.

The most common epileptic syndrome was West Syndrome (WS)(21.25%,68/320), followed by Early-onset Epileptic Encephalopathies (EEEs)(14.06%,45/320), Ohtahara Syndrome (OS)(4.06%,13/320), Dravet Syndrome (DS)(3.44%,11/320), Lennox-Gastaut Syndrome (LGS)(3.13,10/320; Figure S2b).

76.56% (245/320) of patients had comorbidities. For example, global developmental delay was observed in 68.44% of patients (219/320). Brain MRI detected an epileptogenic abnormality such as brain malformations, brain dysplasia, brain atrophy, in 53.13% (170/320) of patients (Table 1).

### Diagnostic Yield

By simultaneously analyzing SNVs and CNVs in the 320 patients, pathogenic or likely pathogenic variants were found in 38.13% (122/320) patients. 104 pathogenic or likely pathogenic SNVs (Table S2) were detected in 28.75%(92/320)patients, 28 pathogenic or likely pathogenic CNVs (Table S3) were found in 8.44%(27/320) patients, concomitant pathogenic SNVs and CNVs were identified 0.63%(2/320) patients and 0 compound heterozygosity with overlapping CNVs and SNVs were found in 31%(1/320) patients (Table 1).

Map of SNVs and CNVs on chromosome idiograms was shown in Figure 1. We found that 24.6% (30/122) of the `pathogenic` or likely pathogenic variants were located on chromosome 16, including 29 SNVs and 6 CNVs. 13.11% (16/122) Of the `pathogenic` or likely pathogenic variants were located on chromosome 2, including 15 SNVs and 2 CNVs.

**Figure 1.**
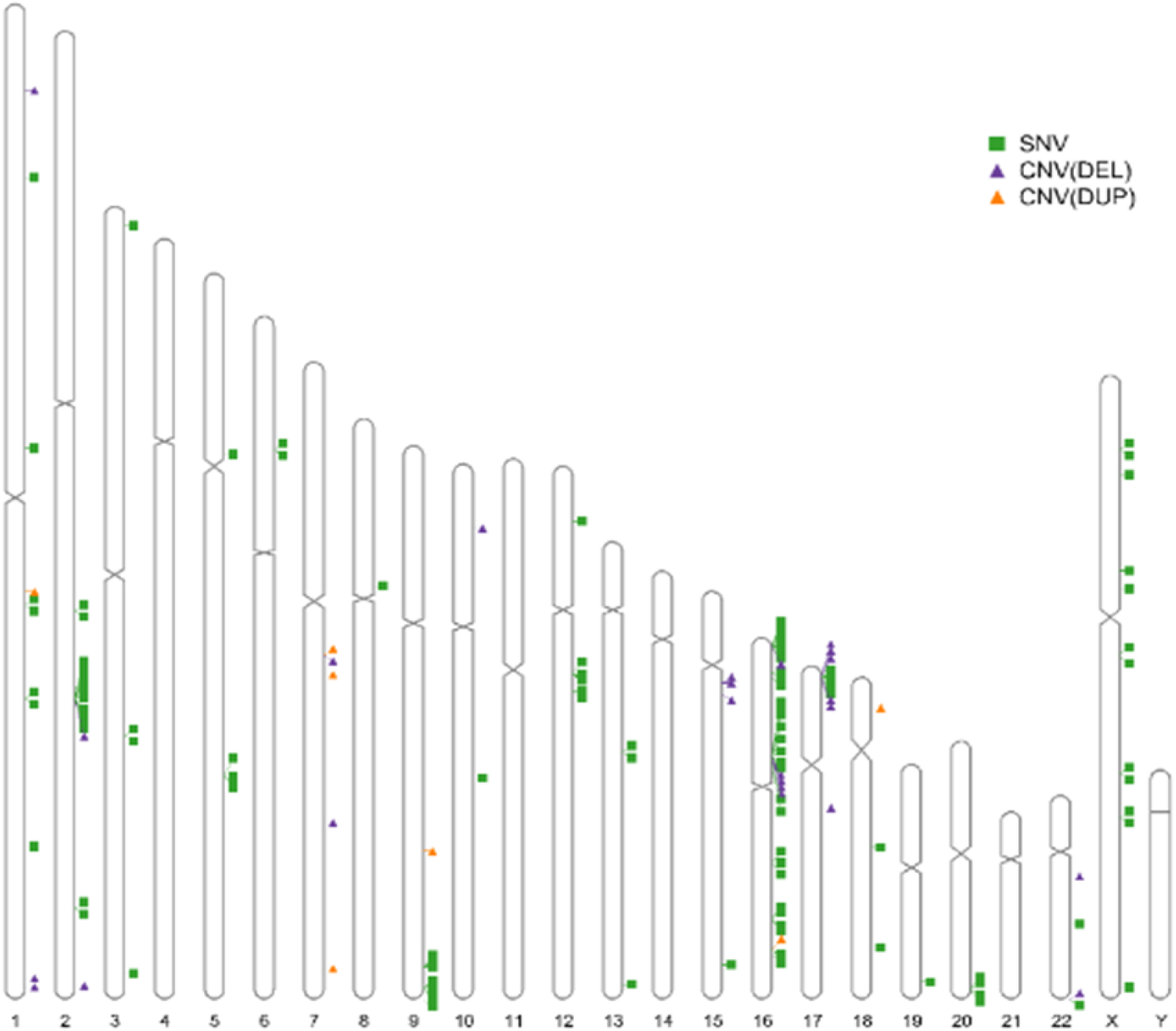
Distribution across chromosomes of associated diagnostic SNVs and diagnostic CNVs. SNVs were marked with green box, and purple triangle represents associated Deletion CNVs, and orange triangle represents associated Duplation CNVs.

233 patients underwent urine screening and tandem mass spectrometry in whole blood. Although no significant metabolic abnormalities were found for these samples, genetic metabolic diseases were detected in 3 percent of patients with normal metabolic screening.

### Factors associated with detection genetic diagnosis

Contrast analysis of the patients with pathogenic or likely pathogenic variants group and the patients without causative variants detection group is shown in Table 1.

Patients with positive result of the genetic detection had a younger average age of seizure onset compared to those with a negative result of the genetic detection [12.84 months vs 21.70 months, p = 0.0003].

Based on the seizure onset age, patients were divided into 6 categories: neonatal, infancy, toddler, early childhood, middle childhood, adolescent. The detection rates were 75%, 38.55%, 33.33%, 24.24%, 10%, 0, respectively. We found that the detection rate decreased with increasing seizure onset age (Figure 2).

**Figure 2.**
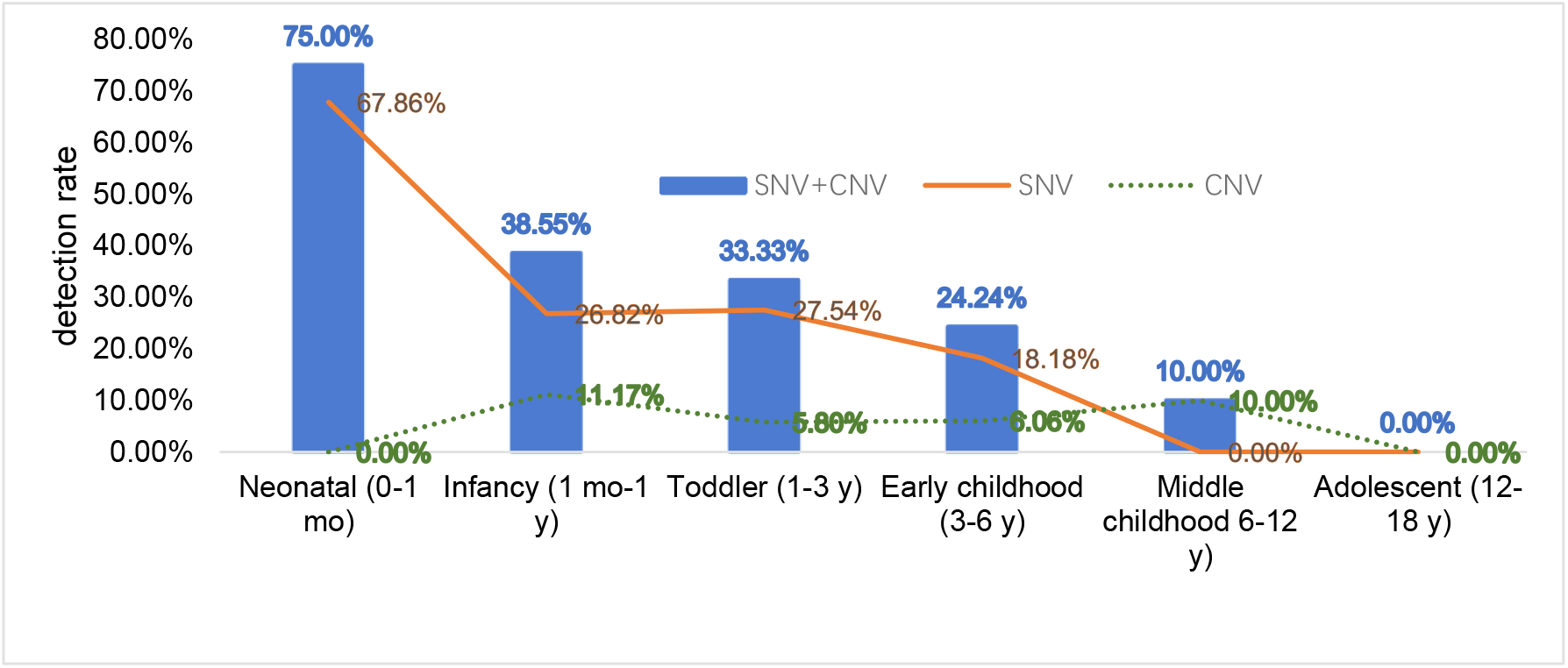
Detection rate of different Age of seizure onset.

Patients with pathogenic or likely pathogenic variants had a younger average age of seizure onset compared to those without an identified genetic etiology [12.84 months vs 21.70 months, p = 0.0003]. There was a statistically significant association between finding a variant and other factor, including developmental delay (7.19E-08), Tuberous Sclerosis Complex (0.003), abnormal limbs (0.028) and Magnetic Resonance Imaging lesions (0.001). We drew a Heatmap of identified genes with diagnostic SNVs and cytoband with diagnostic CNVs among different phenotype subgroups as shown in Figure 3a and 3b.

**Figure 3.**
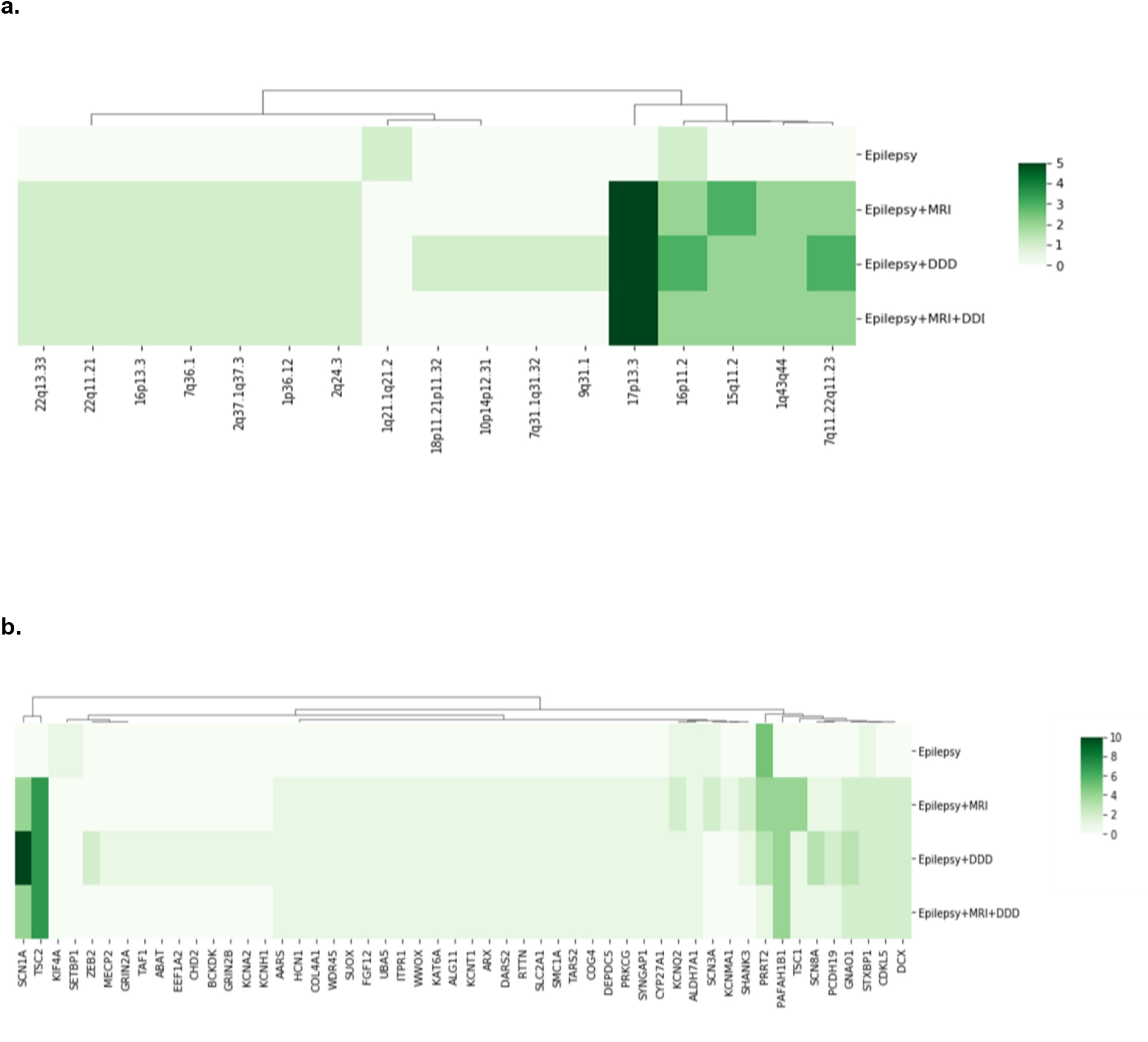
Heatmap of diagnostic CNVs and SNVs among the different phenotype subgroups. Diagnostic CNVs and SNVs of the cohort and the corresponding case subgroups are shown in Figure a and b. DDD, Developmental Delay Disorder MRI, Magnetic Resonance Imaging.

Although there was no significant association between test positive and gender factors(p=0.413), we analyzed a gender-stratified chi-square test of developmental delay and Magnetic Resonance Imaging lesions (Table 2).

**Table 2.** Sex, Development delay and MRI of variants found in epilepsy patients.

|  | Male |  |  | Female |  |  |
| --- | --- | --- | --- | --- | --- | --- |
|  | Positiv | Negative | p | Positiv | Negative | p |
|  | e |  |  | e |  |  |
| **Developmental delay** | 67 | 65 | 4.60E-06 | 37 | 50 | 0.013 |
| **normal development** | 9 | 48 |  | 9 | 35 |  |
| **MRI with finding** | 49 | 53 | 0.113 | 31 | 37 | 0.016 |
| **MRI with no finding** | 23 | 42 |  | 14 | 43 |  |

In the statistical analysis of MRI and positive detection, there was no statistical significance for males(P=0.113, OR=1.688, 95% CI = 0.890-3.201), but had statistical significance for females(P=0.016,OR=2.573,95% CI=1.193- 5.553). In the statistical analysis of developmental delay and positive detection, there was a statistically significant correlation for both males and females.

### Single Nucleotide Variations (SNV)

104 pathogenic or likely pathogenic SNVs in 52 genes were identified in 92 patients (Table S2). The most frequently mutated gene was *SCN1A*(10.87%,10/92), followed by *PRRT2*(8.70%,8/92), *TSC2*(7.61%,7/92), *PAFAH1B1*(4.35%,4/92),*TSC1*(4.35%,4/92),*GNAO1*(3.26%,3/92),*STXBP1*(3.26%,3/92),*KCNQ2*(3.26%,3/92), *SCN8A*(3.26%,3/92), *ZEB2*(2.17%,2/92),*PCDH19*(2.17%,2/92),*CDKL5*(2.17%,2/92),*WWOX*(2.17%,2/92), *ALDH7A1*(2.17%,2/92), *DCX*(2.17%,2/92), *SYNGAP1*(2.17%,2/92), etc.as shown in Table3.

**Table 3.**
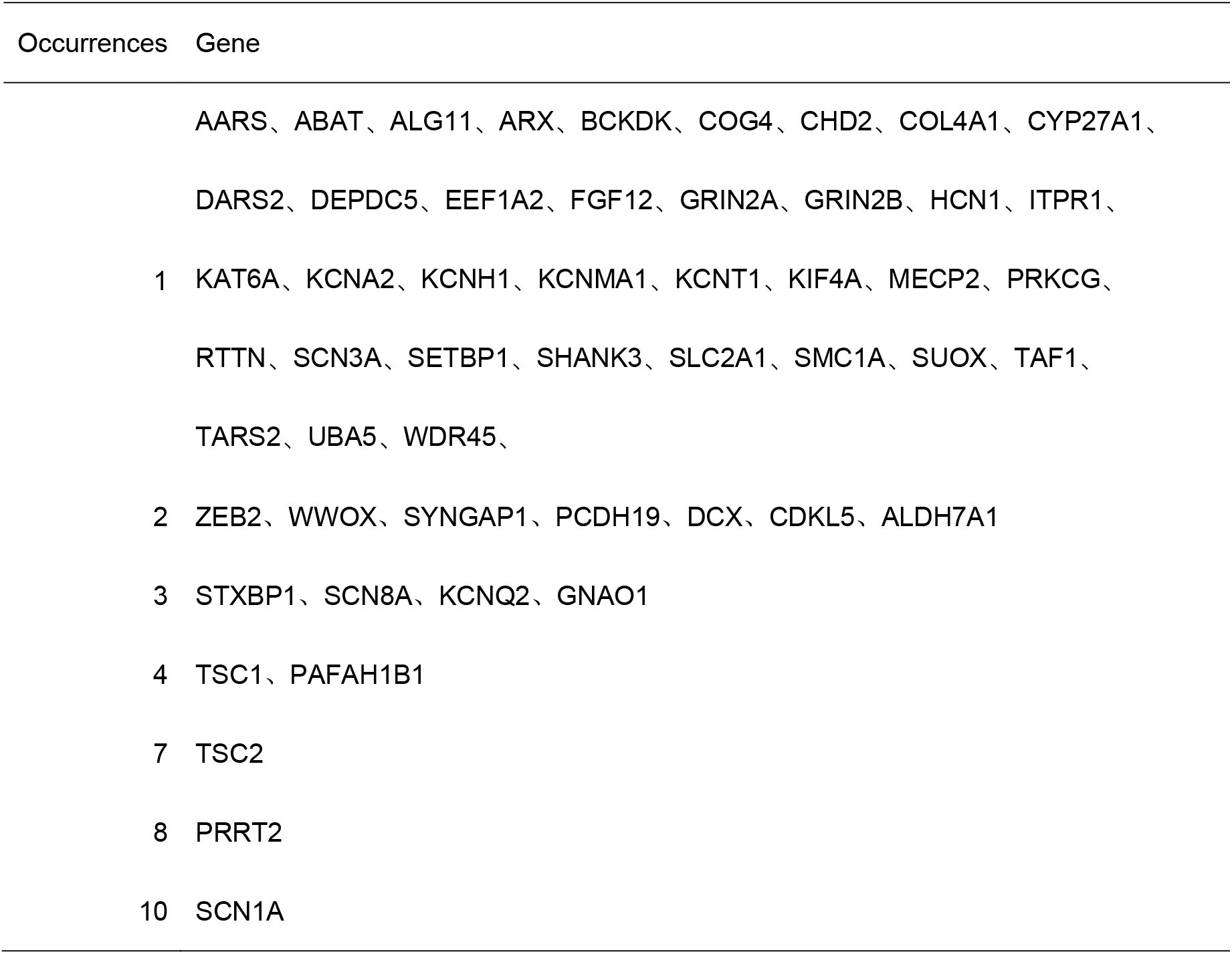
Pathogenic/likely pathogenic genes identified in 320 patients with seizures.

107 SNVs were detected in positive epilepsy patients(Table 5), which involves 52(48.60%)missense variations, 29(27.10%) frameshift variations, 15(14.02%) nonsense variations, 8 (7.48%)splice variations, 1 (0.93%)in frame variations, 1 (0.93%)synonymous variation and 1(0.93%) intron variation. Among them, 61 (57%) were novel variants.

**Table 4.** Variation type in 122 patients with epilepsy.

| **Variation type** | **Number(N, %)** |
| --- | --- |
| **SNV** | **107** |
| Frameshift variation | 29(0.27) |
| Missense variation | 52(0.49) |
| Nonsense variation | 15(0.14) |
| Splicing variation | 8(0.07) |
| in frame insertion | 1(0.01) |
| Synonymous variation | 1(0.01) |
| Intron variation | 1(0.01) |
| **CNV** | **31** |
| Deletion CNV | 24(0.77) |
| Duplation CNV | 7(0.23) |
| <100k CNV | 3(0.10) |
| >100K CNV | 28(0.90) |
| **SNV+CNV** | **3(0.10)** |
| **Total patients** | **122** |

**Table 5.**
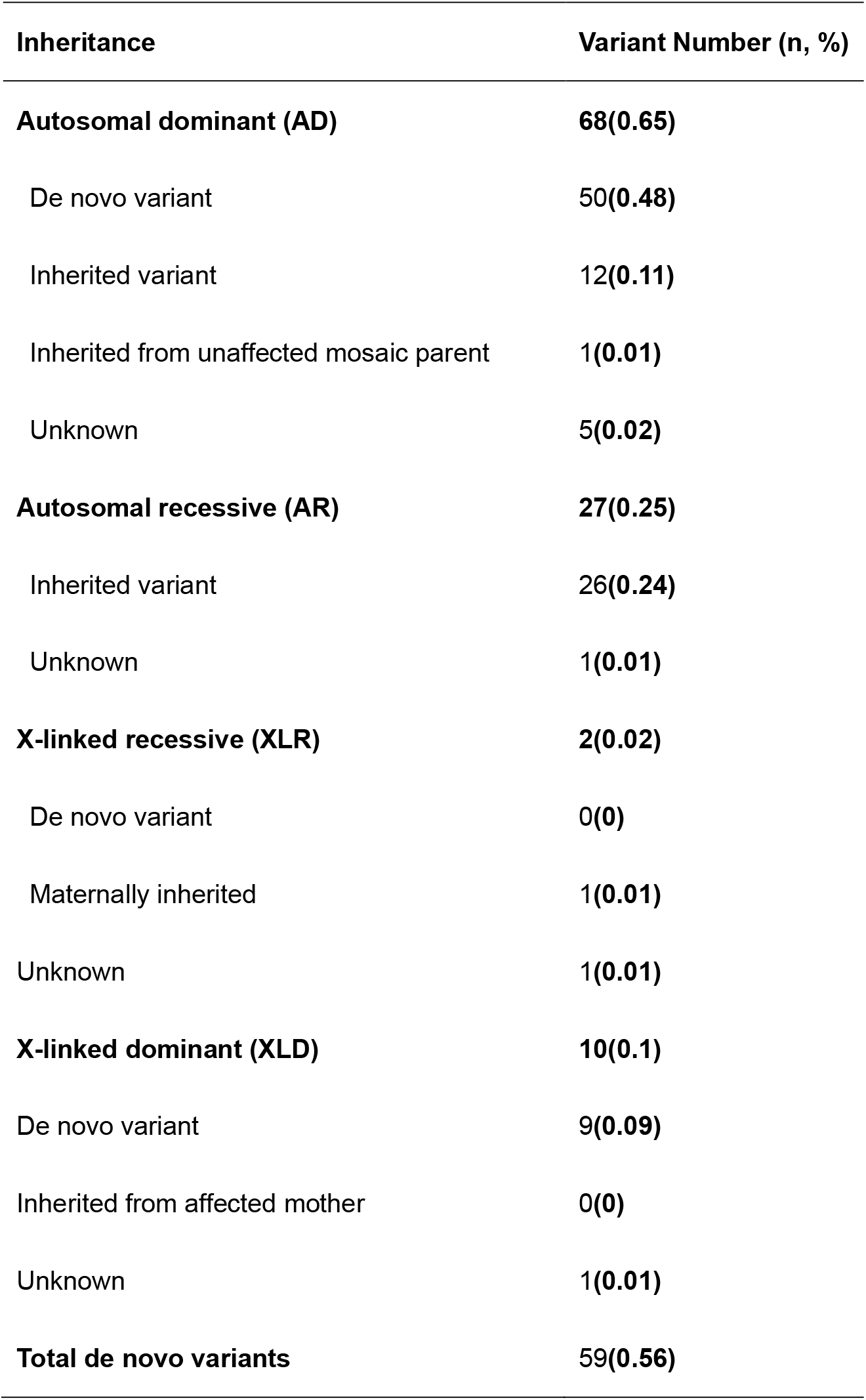
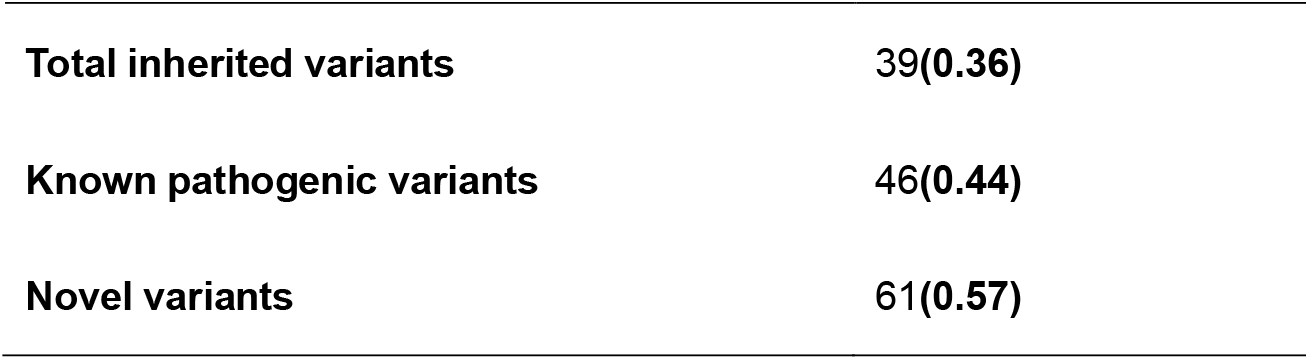
Inheritance patterns among 52 pathogenic /likely pathogenic gene findings from 320 epilepsy patients.

| **Inheritance** | **Variant Number (n, %)** |
| --- | --- |
| **Autosomal dominant (AD)** | **68(0.65)** |
| De novo variant | 50(**0.48**) |
| Inherited variant | 12(**0.11**) |
| Inherited from unaffected mosaic parent | 1(**0.01**) |
| Unknown | 5(**0.02**) |
| **Autosomal recessive (AR)** | **27(0.25)** |
| Inherited variant | 26(**0.24**) |
| Unknown | 1(**0.01**) |
| **X-linked recessive (XLR)** | **2(0.02)** |
| De novo variant | 0(**0**) |
| Maternally inherited | 1(**0.01**) |
| Unknown | 1(**0.01**) |
| **X-linked dominant (XLD)** | **10(0.1)** |
| De novo variant | 9(**0.09**) |
| Inherited from affected mother | 0(**0**) |
| Unknown | 1(**0.01**) |
| **Total de novo variants** | **59(0.56)** |
| **Total inherited variants** | **39(0.36)** |
| **Known pathogenic variants** | **46(0.44)** |
| **Novel variants** | **61(0.57)** |

Inheritance patterns among 52 pathogenic /likely pathogenic gene findings from 320 epilepsy patients are summarized in Table 5, including 68 (71.58%,68/95) autosomal dominant diseases (AD), 15(15.79%, 15/95)autosomal recessive diseases (AR), and 12(12.63%,12/95)X-linked diseases (Table 5, Table S4). Among AD diseases, 73.53% (50/68) variants were de novo (A paternal chimerism sample was included),17.65% (12/68) variants inherited from the parents, 7.35% (5/68) variants were unknown as the samples of the father or mother missed. Among AR diseases, 86.67% (13/15) complex heterozygous variants, 13.33% (2/15) homozygous variants and 16.67% (2/12) X-linked recessive variants were included. Among X-linked diseases,75% (9/12) variants were de novo.

Ten *SCN1A* variants were detected, one variant was inherited from the father, the other 10(90.9%) variants were de novo. Four *TSC1* and 7 *TSC2* variants were all de novo. However, after verification by ddPCR, the father of SZCH0052 had a mosaic variant. Eight *PRRT2* variants were all inherited from the parents, and a frameshift variation(NM_001256442.1: c.640_641insC (p. Arg217Profs8)) was found in 5(3.94%)patients, which was a hotspot of *PRRT2* variant and was incomplete penetrance according to previous studies[27]. Among the epilepsy patients who detected by PRRT2 variants, the types of variants were all nonsense variants or frameshift variants. Six variants occur at the codon 217, and the age of seizures onset in this six samples were less than 1 year old.

One patient had a complex heterozygous variant of the *BCKDK* gene. The gene could cause the Branched- chain ketoacid dehydrogenase kinase deficiency (MIM#614923) in OMIM and with no specific genetic pattern. The seizure onset of the patient was at 6 years old, who had neurodevelopmental disorders and autism. Our results obtained incriminate *BCKDK* as a gene for recessive Branched-chain ketoacid dehydrogenase kinase deficiency (MIM#614923).

### Copy number variants (CNV)

Thirty-one diagnostic CNVs were identified in 30 patients(SZCH0370 had two CNVs)ranging from 121 bp to 30.47Mb. 77.42% (24/71) copy number deletions and 22.58% (7/31) copy number duplications were found in our cohort. According to our interpretation rules (Figure S1), among these variants, 80.65% (25/31) CNVs were pathogenic, 19.35% (6/31) CNVs were likely pathogenic.

The CNVs were located on 12 chromosomes shown in Figure 2. The most common syndrome was Chromosome 17p13.3 deletion syndrome (1.53%,5/320), also known as Miller-Dieker lissencephaly syndrome (MIM#247200), which could result in deletion of the *PAFAH1B1* gene. Deletion or mutation in the *PAFAH1B1* gene may lead to lissencephaly (OMIM#607432). We detected 4 pathogenic/likely pathogenic SNVs in the other 4 patients with lissencephaly. In our research 16 patients developed lissencephaly, 9 of whom had *PAFAH1B1* gene variants, accounting for 56.25% (9/16) of the causes of lissencephaly (Table S5). In patients with *PAFAH1B1* gene variants usually demonstrated more severe lissencephaly in the parietal-occipital region of the brain and the severity of the malformation increased from anterior to posterior head.

The other syndromes caused by recurrent CNVs were Chromosome 16p11.2 deletion syndrome (OMIM #611913, 4/320), Chromosome 15q11.2 deletion syndrome (MIM#615656, 3/320) and Chromosome 7q11.23 duplication syndrome (MIM#609757, 2/320) as shown in Table S3.

In our study, most of the diseases caused by chromosomal abnormalities were known syndromes (Table S3). Seizure was one of the phenotypes, such as Chromosome 16p11.2 deletion syndrome. Some other CNVs may cover epileptic pathogenic genes, and the copy number deletion or duplication leads to epilepsy in patients, such as: a 1.5-Mb deletion at 2q24.3 in SZCH0255, covering 9 genes: *SCN1A, XIRP2, TTC21B, SCN9A, SCN7A, CSRNP3, LOC100506134, LOC100506124, GALNT3*. The deletions of *SCN1A* and *SCN9A* cause epilepsy.

Among the 31 CNVs, the length of 3 CNVs was below 25K. According to the detection technique of current CMA, CNV fragments below 25K cannot be detected[28]. However, the detection of small fragments of CNV is a major advantage of GS. The rate of genetic diagnosis of epilepsy has improved 1% (3/320).

We detected a 1651bp deletion in 16p13.3. Two exons of TSC2 were deleted in the CNV. To verify our detection technology for CNVs, we validated the CNV in the SZCH0053 sample. By designing primers outside the CNV breakpoint, we predicted that if the CNV exists, a 237bp product will be obtained. The 237-bp product and a heterozygous band of normal fragments was found in the patient, and the CNV was not detected in his healthy parents (Figure 4). The results were shown in the figure, demonstrating that the CNV is de novo.

**Figure 4.**
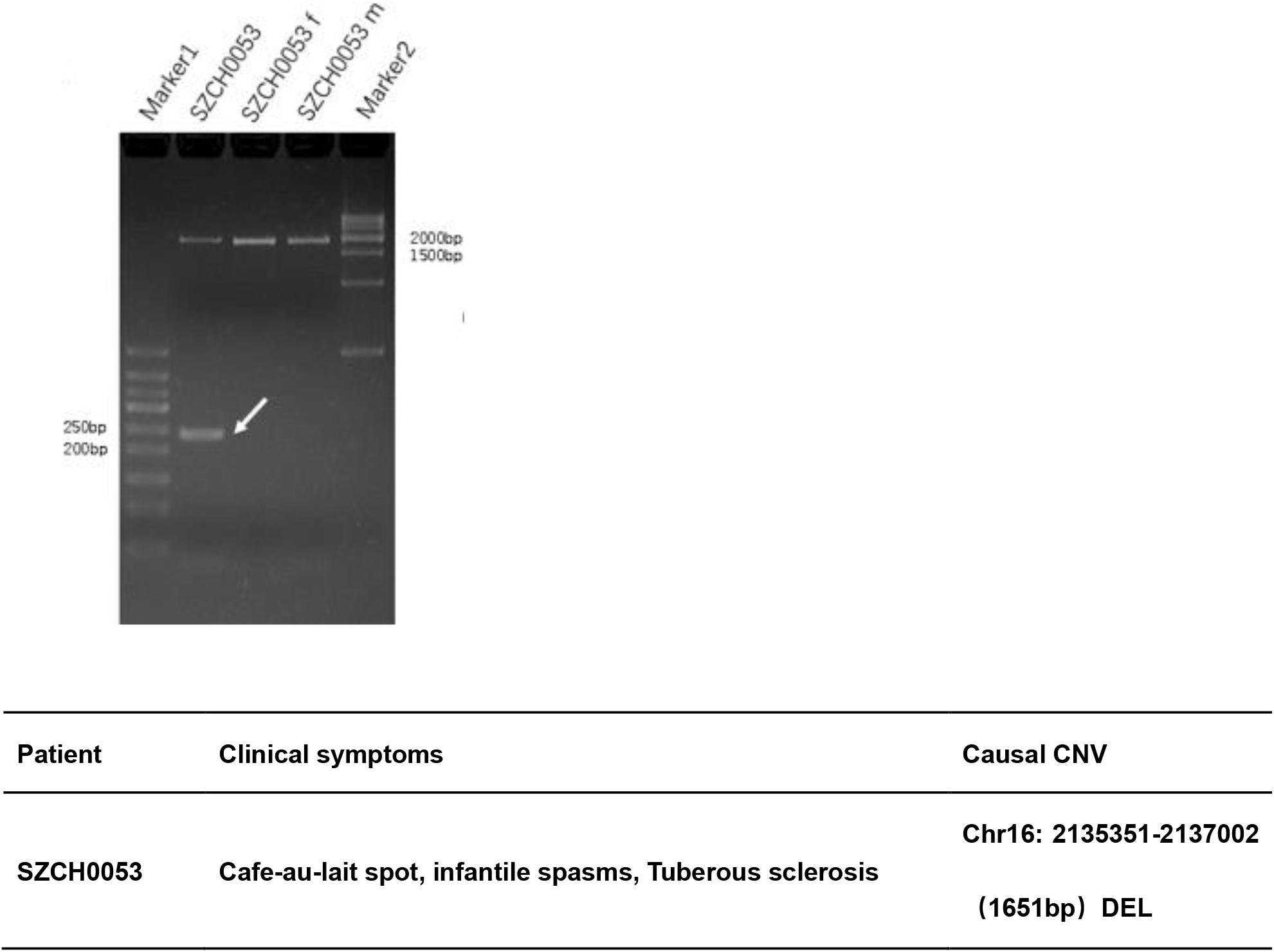
A de novo heterozygous deletion CNV of the TSC2 in SZCH053.

### Cases diagnosed by combinational CNV and SNV analysis

We also found that a 2-year-old girl (SZCH0243) harbored concomitantly pathogenic SNV and a pathogenic CNV. She presented with seizures on the first day after birth, hypotonia, mental retardation, carrying a *KCNQ2* (NM_004518.5: c.953T>C (p. Leu318Pro)) variant and a 684kb deletion in 15q11.2. In this sample, the *KCNQ2* variant was *de novo*, which can result in Epileptic encephalopathy, early infantile (OMIM#613720). The CNV-causing disease was Chromosome 15q11.2 deletion syndrome. This was considered dual-diagnosed case.

A 1-year-old girl (SZCH0677) also harbored concomitantly a pathogenic SNV and a pathogenic CNV. She presented with seizures at ten months of age, mental retardation, carrying a *SYNGAP1* (NM_001130066.1: c.1543C>T (p. Arg515Cys)) variant and a 1542kb deletion in 17q12. In this sample, the *SYNGAP1* variant was *de novo*, which can result in *SYNGAP1* (OMIM#612621). The CNV-causing disease was Chromosome 17q12 deletion syndrome. This was considered dual-diagnosed case.

A 2-year-old boy (SZCH0268) had a SNV and intragenic CNV. He presented with seizures in the first month of life, hypertonia, mental retardation and corpus callosum dysplasia. We detected a splice variant (NM_001291997.1: c.453-1G>C) on the WWOX gene and a CNV duplication, which resulted in partial duplication of exon6-exon8. However, we could not confirm whether this SNV and CNV could form a compounded heterozygosity due to because of experimental reasons WWOX gene (MIM*605131) was associated with Epileptic encephalopathy, early infantile (OMIM#616211), AR inheritance. The SNV and CNV complex heterozygous disease on WWOX has been reported in 2015[29]. We supposed that these two variants which overlapped on the same gene and formed homozygous variant were the cause of the disease.

## EFFECT ON CLINICAL MANAGEMENT

At present, the treatment for epilepsy is mainly aimed at controlling seizures. Treatment selections are made according to the seizure types and electro-clinical syndromes.

GS can sequence the entire genome of an individual, not only for disease diagnosis in patients with epilepsy, but also for drug selection. 43 patients (13.15%) were diagnosed with treatable disorders according to molecular diagnosis, two pyridoxine-dependent epilepsy and one GLUT1 deficiency syndrome was confirmed. In addition, 3 patients with hereditary metabolic diseases received direct therapeutic measures and demonstrated favorable prognosis as shown in Table 6.

**Table 6.**
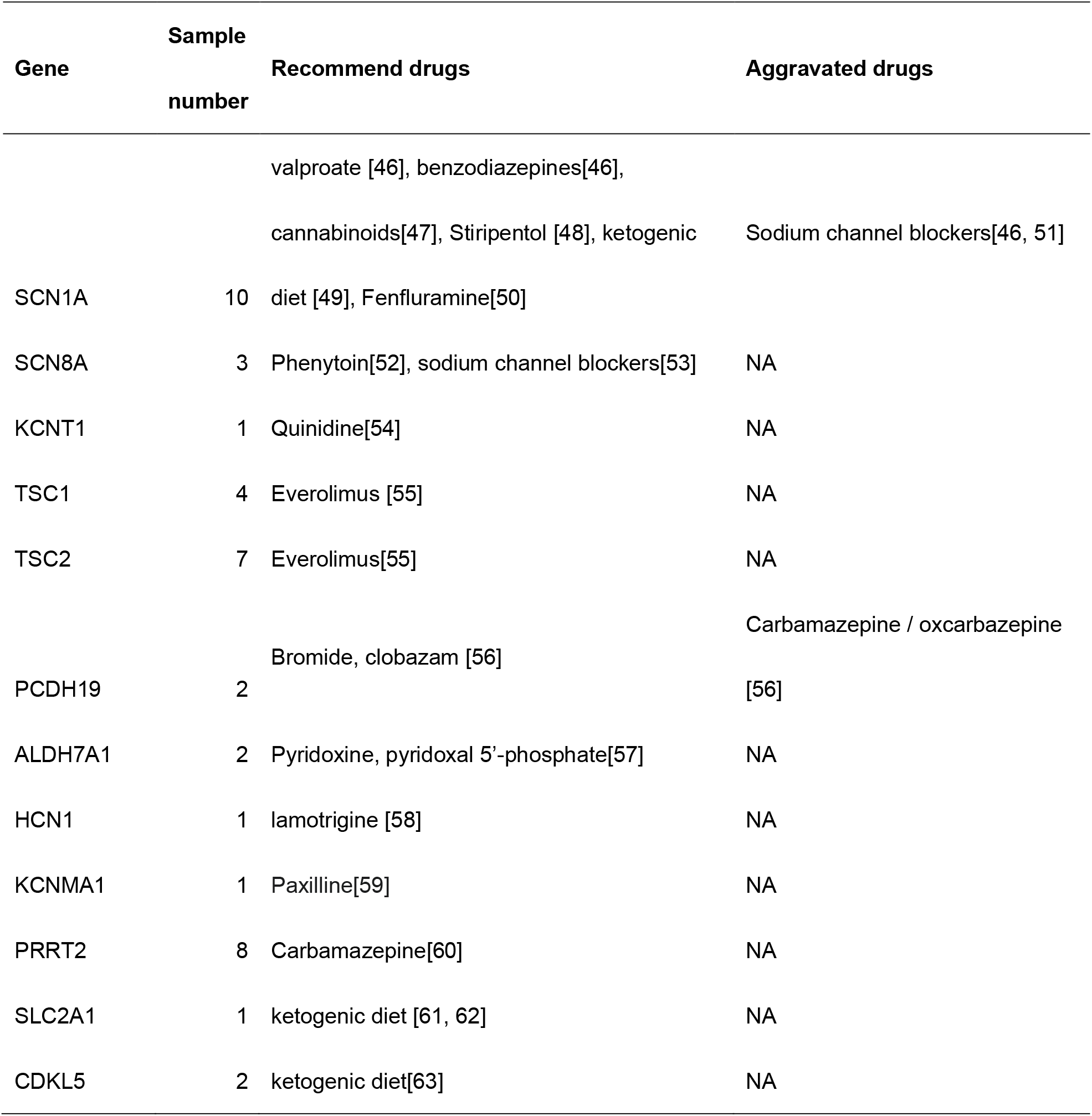
Gene variants associated with appropriate treatment regimens.

An eight-month-old girl (SZCH0292) developed seizures on the 13th day after birth and the seizure was refractory after taking several antiepileptic drugs. We detected two complex heterozygous variants on the ALDH7A1 gene (OMIM#107323) (NM_001182.4: c.1547A>G (p. Tyr516Cys) and NM_001182.4: c.965C>T (p. Ala322Val)). The girl achieved seizure free and had a good prognosis after she was treated with VitB6.

A three-year-old boy (SZCH0283) had seizures at three months old, with ataxia and mental retardation. We detected a heterozygous frameshift variant on SLC2A1 (OMIM*138140) (NM_006516.2: c.912del (p. Gln304Hisfs36)). The boy achieved seizure free after ketogenic diet, normal intellectual and motor development.

### Recurrence risk

Among the 92 patients with pathogenic/likely pathogenic SNVs, 42.39% (39/92) patients inherited the variant from their parents. This result suggests that in these families, there is a risk of recurrence in other family members.

Among 59 patients with de novo variants, only 3 families (5.08%) had two children with the same birth defects, indicating that the probability of having two or more children with the same defective problem was low in this circumstance. If the patient’s pathogenic site is a de novo variant, there is theoretically no risk of recurrence for siblings in the family. However, parental mosaicism increases the risk of recurrence and might be more prevalent than previously predicted [5]. In our study, we found a patient’s father had a chimeric TSC1 variant. In this case, there is a certain risk of recurrence in fertility.

## Discussion

We conducted GS on 320 children with epilepsy. The diagnostic yield was 38.13% (122/320), in which CNV was 8.44% (27/320).

There are few studies about conducting GS on such a large-scale sample of pediatric patients. Most of the current large-scale studies underwent ES or capture-based targeted sequencing[30-33].

The diagnostic yield of previous studies on epilepsy was about 18%-49%[34-38]. The patients with epilepsy in different studies were different, and the filtering and interpretation methods for the variants were different. Therefore, it was difficult to compare the diagnostic yield among different studies.

Based on the current results of a genetic test for 320 patients with epilepsy by GS, if we use other methods for testing, we compare the possible results between ES or capture-based targeted sequencing as shown in Figure 5. The increased diagnostic yield in the GS was due to the detection of CNVs and intronic SNVs.

**Figure 5.**
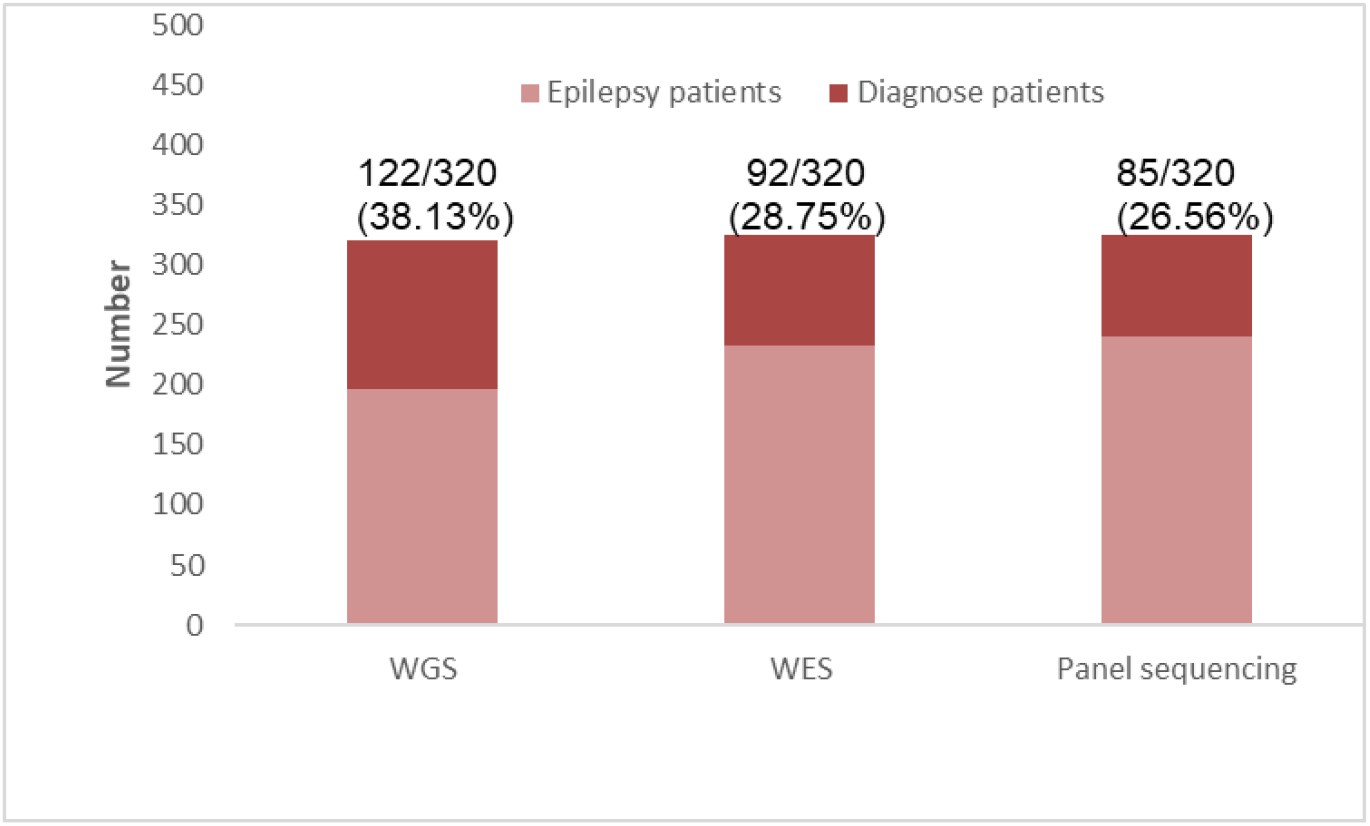
Comparison between GS, ES and capture-based targeted sequencing in detection of 320 epilepsy patients. Capture-based targeted sequencing: referring to an epilepsy panel that includes a total of 813 genes from MNG Laboratories (Atlanta, GA, USA)[40]

GS can not only sequence for SNVs and small fragment insertion, but also analyze CNVs, which substitute for individual chromosome analysis. Further GS is superior to CMA in terms of the identifiable size of CNV [39]. In our study, 3 CNVs were detected below 25K, which could not be detected by CMA. Compared with the CMA and ES, the detection rate is improved at least 1%.

GS sequencing can cover the entire UTR and intron regions, while ES can only detect the coding region of genes. Although currently the functionality of most UTR and intron regions is unknown, ES can missed detection as pathogenic or functional areas associated with epilepsy are constantly being discovered with the development of non-coding regions, such as the 20N region of SCN1A[7].

Compared with capture-based targeted sequencing, the advantages of GS were more obvious. Referring to a panel for epilepsy that includes 813 genes from MNG Laboratories (Atlanta, GA, USA)[40], 15.69%(8/51)genes detected in our study were not included in this panel. GS increased the detection rate at least 11.88%.

Our study found that patients who developed seizures within one month after birth had a genetic diagnosis rate of 75% (21/28). As the age of seizure onset increased, the diagnostic yield decreased, indicating that the earlier the patients develop epilepsy, the higher possibility the seizures are caused by genetic factors[30]. Our diagnostic yield exceeded other previous studies in this seizure onset age group[41, 42]. Two patients in this group of positive diagnostics had both CNV and SNV pathogenic variants, indicating that a traditional single CNV or SNV test were easily missed detection, which proves the necessity of simultaneous detection of SNV and CNV in the epileptic seizure patients of neonatal. GS can simultaneously analyze the SNV and CNV of the individual, showing that GS can be performed in the epileptic seizure patients of neonatal, which can more accurately perform early diagnosis and early intervention and early treatment.

In our study, patients with epilepsy and other phenotypes accounted for 76.56%, but genes potentially associated with various phenotypes were not necessarily included in the list of genes that were clearly associated with each phenotype. For example, seizure was only one of the phenotypes in some patients, and may not be a significant feature of the patient. In this circumstance, capture-based targeted sequencing might miss the detection. Similarly, multiple phenotypes demonstrated in an individual may be caused by different variants in different genes, or a heterozygous CNV and SNV combination, such as SZCH0268.

Such results illustrated the importance of simultaneous analysis of both SNVs and CNVs. Compared with traditional analysis, it could also provide a more comprehensive genetic interpretation for the genetic diagnosis of diseases. In our study, we also analyzed repeat expansions given the WGS platform but we did not get the positive results.

We found that the average age of seizure onset in the diagnostic SNV patients (11.58 month) was young than that of the CNV-caused patients (18.15 month). We also compared the relationship between CNV length and age of seizure onset as shown in Figure S3, and found that the longer the CNV fragment, the older the average age of seizure onset. In SZCH0370 and SZCH0053, which with CNV length smaller than 100K, copy number variation resulted in the premature termination of protein synthesis and the age of seizure onset in both patients was less than 1 year old. Therefore, the sooner or later of the age of seizure onset may be related to the degree of variation of epilepsy related genes.

We can completely save all the genetic data of the individual by performing GS on patients with epilepsy only once, without sampling and sequencing again. After the discovery of some new genes and variants, we can return to the original GS data for interpretation. However, ES and capture-based targeted sequencing do not have such advantages. The second test will increase the cost, and some samples may not be obtained again due to unexpected factors, such as missing or dead. GS retains all the genetic information for individual genetic diagnosis. Sustainability of the research has prominent significance for patients’ family reproduction.

. An important purpose of genetic diagnosis is to provide targeted clinical management of patients with epilepsy. At present, personalized treatment has become possible. We can not only accurately use the genetic diagnosis results of patients with epilepsy such as Pyridoxine-dependent seizures and GLUT1 deficiency syndrome 1, but also conduct research related to pharmacokinetics.

Two pyridoxine-dependent epilepsy and one GLUT1 deficiency syndrome was confirmed in our study. The patients started to have seizure onset when they were early infantile. After we performed proper treatment on them, the prognosis is prominent, they achieved seizure free and demonstrated normal development. It was important to conduct early detection and initiate early treatment for patients with early infantile epilepsy, to improve the prognosis and outcome. It would also affect further treatment strategies. It has a strong predictive value for long-term cognitive and developmental outcome.[43, 44].

Testing GS can achieve the genetic diagnosis, individualized treatment, and family reproductive guidance, which can reduce the health care cost of patients with epilepsy. Research has shown that performing ES on the patients at the first time for diagnosis is the most cost-effective test for epilepsy[45]. As sequencing costs continue to decline, we believe that GS can replace ES for the genetic testing for patients with epilepsy as GS can provide more information and have a higher detective rate than ES.

In summary, GS has advantages in the genetic diagnosis for patients with epilepsy, especially for patients with neonatal seizures. Early GS testing can provide an accurate molecular diagnosis in a timely manner, predict possible phenotypes, initiate appropriate follow-up, predict prognosis, and assess the risk of recurrence, which is helpful for the clinical management of these patients.

By performing trios-based GS, the diagnostic yield can be improved and the assessment for recurrence risk of the family can be more accurate. In future studies, we plan to conduct GS on `patients` without causative variants in our study.

## Data Availability

The data that support the findings of this study have been deposited in the CNSA (https://db.cngb.org/cnsa/) of CNGBdb with accession code CNP0000788.

## Acknowledgement

This research was supported by Science, Technology and Innovation Commission of Shenzhen Municipality under grant No. KJYY20151116165726645.

All sequencing data was produced by China National GeneBank, Shenzhen, China. The data that support the findings of this study have been deposited in the CNSA (https://db.cngb.org/cnsa/) of CNGBdb with accession code CNP0000788.

## REFERENCE

1. Neligan, A., W.A. Hauser, and J.W. Sander, The epidemiology of the epilepsies. Handb Clin Neurol, 2012. 107: p. 113–33.

2. Weber, Y.G., et al., The role of genetic testing in epilepsy diagnosis and management. Expert Rev Mol Diagn, 2017. 17(8): p. 739–750.

3. Steinlein, O.K., et al., A missense mutation in the neuronal nicotinic acetylcholine receptor alpha 4 subunit is associated with autosomal dominant nocturnal frontal lobe epilepsy. Nat Genet, 1995. 11(2): p. 201–3.

4. Wang, J., et al., Epilepsy-associated genes. Seizure, 2017. 44: p. 11–20.

5. Dunn, P., et al., Next Generation Sequencing Methods for Diagnosis of Epilepsy Syndromes. Front Genet, 2018. 9: p. 20.

6. Stavropoulos, D.J., et al., Whole-genome sequencing expands diagnostic utility and improves clinical management in paediatric medicine. NPJ genomic medicine, 2016. 1(1): p. 1–9.

7. Carvill, G.L., et al., Aberrant Inclusion of a Poison Exon Causes Dravet Syndrome and Related SCN1A-Associated Genetic Epilepsies. Am J Hum Genet, 2018. 103(6): p. 1022–1029.

8. Corbett, M.A., et al., Intronic ATTTC repeat expansions in STARD7 in familial adult myoclonic epilepsy linked to chromosome 2. Nat Commun, 2019. 10(1): p. 4920.

9. Meienberg, J., et al., New insights into the performance of human whole-exome capture platforms. Nucleic Acids Res, 2015. 43(11): p. e76.

10. Fisher, R.S., et al., Operational classification of seizure types by the International League Against Epilepsy: Position Paper of the ILAE Commission for Classification and Terminology. Epilepsia, 2017. 58(4): p. 522–530.

11. Huang, J., et al., A reference human genome dataset of the BGISEQ-500 sequencer. Gigascience, 2017. 6(5): p. 1–9.

12. Goyal, A., et al., Ultra-fast next generation human genome sequencing data processing using DRAGENTM bio-IT processor for precision medicine. 2017.

13. Chiang, C., et al., SpeedSeq: ultra-fast personal genome analysis and interpretation. Nat Methods, 2015. 12(10): p. 966–8.

14. Layer, R.M., et al., LUMPY: a probabilistic framework for structural variant discovery. Genome Biol, 2014. 15(6): p. R84.

15. Dong, Z., et al., Low-pass whole-genome sequencing in clinical cytogenetics: a validated approach. Genet Med, 2016. 18(9): p. 940–8.

16. Lek, M., et al., Analysis of protein-coding genetic variation in 60,706 humans. Nature, 2016. 536(7616): p. 285.

17. Karczewski, K.J., et al., Variation across 141,456 human exomes and genomes reveals the spectrum of loss-of-function intolerance across human protein-coding genes. BioRxiv, 2019: p. 531210.

18. Genomes Project, C., et al., A global reference for human genetic variation. Nature, 2015. 526(7571): p. 68–74.

19. Landrum, M.J., et al., ClinVar: public archive of interpretations of clinically relevant variants. Nucleic Acids Res, 2016. 44(D1): p. D862–8.

20. Solomon, B.D., et al., Clinical genomic database. Proc Natl Acad Sci U S A, 2013. 110(24): p. 9851–5.

21. Hamosh, A., et al., Online Mendelian Inheritance in Man (OMIM), a knowledgebase of human genes and genetic disorders. Nucleic Acids Res, 2005. 33(Database issue): p. D514–7.

22. Stenson, P.D., et al., Human gene mutation database (HGMD®): 2003 update. Human mutation, 2003. 21(6): p. 577–581.

23. Richards, S., et al., Standards and guidelines for the interpretation of sequence variants: a joint consensus recommendation of the American College of Medical Genetics and Genomics and the Association for Molecular Pathology. Genetics in medicine, 2015. 17(5): p. 405.

24. Riggs, E.R., et al., Technical standards for the interpretation and reporting of constitutional copy-number variants: a joint consensus recommendation of the American College of Medical Genetics and Genomics (ACMG) and the Clinical Genome Resource (ClinGen). Genet Med, 2019.

25. Richards, S., et al., Standards and guidelines for the interpretation of sequence variants: a joint consensus recommendation of the American College of Medical Genetics and Genomics and the Association for Molecular Pathology. Genet Med, 2015. 17(5): p. 405–24.

26. Hao, Z., et al., RIdeogram: drawing SVG graphics to visualize and map genome-wide data on the idiograms. PeerJ Computer Science, 2020. 6: p. e251.

27. Meneret, A., et al., PRRT2 mutations: a major cause of paroxysmal kinesigenic dyskinesia in the European population. Neurology, 2012. 79(2): p. 170–4.

28. Fish, M., Analysis of genetic variations associated with arrhythmogenic right ventricular cardiomyopathy. 2016, University of Cape Town.

29. Mignot, C., et al., WWOX-related encephalopathies: delineation of the phenotypical spectrum and emerging genotype-phenotype correlation. J Med Genet, 2015. 52(1): p. 61–70.

30. Demos, M., et al., Diagnostic Yield and Treatment Impact of Targeted Exome Sequencing in Early-Onset Epilepsy. Front Neurol, 2019. 10: p. 434.

31. Epi25 Collaborative. Electronic address, s.b.u.e.a. and C. Epi, Ultra-Rare Genetic Variation in the Epilepsies: A Whole-Exome Sequencing Study of 17,606 Individuals. Am J Hum Genet, 2019. 105(2): p. 267–282.

32. Costain, G., et al., Clinical Application of Targeted Next-Generation Sequencing Panels and Whole Exome Sequencing in Childhood Epilepsy. Neuroscience, 2019. 418: p. 291–310.

33. Truty, R., et al., Possible precision medicine implications from genetic testing using combined detection of sequence and intragenic copy number variants in a large cohort with childhood epilepsy. Epilepsia Open, 2019. 4(3): p. 397–408.

34. Butler, K.M., et al., Diagnostic Yield From 339 Epilepsy Patients Screened on a Clinical Gene Panel. Pediatr Neurol, 2017. 77: p. 61–66.

35. Helbig, K.L., et al., Diagnostic exome sequencing provides a molecular diagnosis for a significant proportion of patients with epilepsy. Genetics in Medicine, 2016. 18(9): p. 898.

36. Yang, L., et al., Clinical and genetic spectrum of a large cohort of children with epilepsy in China. Genet Med, 2019. 21(3): p. 564–571.

37. Tumiene, B., et al., Diagnostic exome sequencing of syndromic epilepsy patients in clinical practice. Clinical genetics, 2018. 93(5): p. 1057–1062.

38. Liang, J.-S., et al., Genetic diagnosis in children with epilepsy and developmental delay/mental retardation using targeted gene panel analysis. 2018.

39. Zhou, B., et al., Whole-genome sequencing analysis of CNV using low-coverage and paired-end strategies is efficient and outperforms array-based CNV analysis. J Med Genet, 2018. 55(11): p. 735–743.

40. Baldassari, S., et al., Dissecting the genetic basis of focal cortical dysplasia: a large cohort study. Acta Neuropathol, 2019.

41. Jang, S.S., et al., Diagnostic Yield of Epilepsy Panel Testing in Patients With Seizure Onset Within the First Year of Life. Front Neurol, 2019. 10: p. 988.

42. Olson, H.E., et al., Genetics and genotype-phenotype correlations in early onset epileptic encephalopathy with burst suppression. Ann Neurol, 2017. 81(3): p. 419–429.

43. Mizrahi, E.M. and R.R. Clancy, Neonatal seizures: Early-onset seizure syndromes and their consequences for development. Mental retardation and developmental disabilities research reviews, 2000. 6(4): p. 229–241.

44. Tadic, B.V., et al., Long-term outcome in children with neonatal seizures: A tertiary center experience in cohort of 168 patients. Epilepsy & Behavior, 2018. 84: p. 107–113.

45. Sanchez Fernandez, I., et al., Diagnostic yield of genetic tests in epilepsy: A meta-analysis and cost-effectiveness study. Neurology, 2019.

46. Wirrell, E.C., et al., Optimizing the diagnosis and management of Dravet syndrome: recommendations from a North American consensus panel. Pediatric neurology, 2017. 68: p. 18-34. e3.

47. Devinsky, O., et al., Trial of cannabidiol for drug-resistant seizures in the Dravet syndrome. New England Journal of Medicine, 2017. 376(21): p. 2011–2020.

48. Chiron, C., et al., Stiripentol in severe myoclonic epilepsy in infancy: a randomised placebo-controlled syndrome-dedicated trial. The Lancet, 2000. 356(9242): p. 1638–1642.

49. Ko, A., et al., The efficacy of ketogenic diet for specific genetic mutation in developmental and epileptic encephalopathy. Frontiers in Neurology, 2018. 9: p. 530.

50. Bialer, M., et al., Progress report on new antiepileptic drugs: a summary of the Fourteenth Eilat Conference on new antiepileptic drugs and devices (EILAT XIV). I. Drugs in preclinical and early clinical development. Epilepsia, 2018. 59(10): p. 1811–1841.

51. Shi, X.-Y., et al., Efficacy of antiepileptic drugs for the treatment of Dravet syndrome with different genotypes. Brain and Development, 2016. 38(1): p. 40–46.

52. Boerma, R.S., et al., Remarkable phenytoin sensitivity in 4 children with SCN8A-related epilepsy: a molecular neuropharmacological approach. Neurotherapeutics, 2016. 13(1): p. 192–197.

53. Møller, R.S. and K.M. Johannesen, Precision medicine: SCN8A encephalopathy treated with sodium channel blockers. Neurotherapeutics, 2016. 13(1): p. 190–191.

54. Bearden, D., et al., Targeted treatment of migrating partial seizures of infancy with quinidine. Annals of neurology, 2014. 76(3): p. 457–461.

55. Franz, D.N., et al., Everolimus for treatment-refractory seizures in TSC: Extension of a randomized controlled trial. Neurology: Clinical Practice, 2018. 8(5): p. 412–420.

56. Lotte, J., et al., Effectiveness of antiepileptic therapy in patients with PCDH19 mutations. Seizure, 2016. 35: p. 106–110.

57. Mills, P.B., et al., Mutations in antiquitin in individuals with pyridoxine-dependent seizures. Nature medicine, 2006. 12(3): p. 307.

58. Reid, C.A., A.M. Phillips, and S. Petrou, HCN channelopathies: pathophysiology in genetic epilepsy and therapeutic implications. British journal of pharmacology, 2012. 165(1): p. 49–56.

59. Sheehan, J.J., B.L. Benedetti, and A.L. Barth, Anticonvulsant effects of the BK-channel antagonist paxilline. Epilepsia, 2009. 50(4): p. 711–720.

60. Huang, X.-J., et al., Paroxysmal kinesigenic dyskinesia: Clinical and genetic analyses of 110 patients. Neurology, 2015. 85(18): p. 1546–1553.

61. Klepper, J., et al., Seizure control and acceptance of the ketogenic diet in GLUT1 deficiency syndrome: a 2-to 5-year follow-up of 15 children enrolled prospectively. Neuropediatrics, 2005. 36(05): p. 302–308.

62. Klepper, J., et al., Effects of the ketogenic diet in the glucose transporter 1 deficiency syndrome. Prostaglandins, leukotrienes and essential fatty acids, 2004. 70(3): p. 321–327.

63. Lim, Z., et al., Use of the ketogenic diet to manage refractory epilepsy in CDKL 5 disorder: Experience of> 100 patients. Epilepsia, 2017. 58(8): p. 1415–1422.

